# Defining the Components of a Nurse Practitioner-Led Home Visit Intervention for Frail Patients: An International Delphi Consensus Study

**DOI:** 10.64898/2026.08.14.26360429

**Authors:** Ariane Sacchetti, Alexandre Bellier, Christophe Pison, Mélanie Berube

## Abstract

**Purpose:** Aging in place has become a central objective of health and social policies across the world, yet frailty and multimorbidity significantly undermine individuals’ capacity to remain safely at home. The aim was to identify the potential components of a home visit intervention led by nurse practitioners for frail populations.

**Design:** A consensus study using a two-round Delphi method

**Methods:** A two-round Delphi study was conducted in summer 2024 with 15 experts from four French-speaking countries (Quebec–Canada, Switzerland, Belgium, and France). The questionnaire was based on documented needs of frail patients and their caregivers.

**Results:** Experts identified the target population as older adults needing home care, people with physical or cognitive impairments, those requiring end-of-life care, and individuals experiencing difficulties remaining at home. Eligibility criteria included frailty, multiple chronic conditions, mobility issues, social isolation, and low socio-economic status. The nurse practitioner’s role should include clinical assessment, treatment adjustments, care coordination, therapeutic education, support for patients and families, and promotion of self-care. Nurse practitioners may also serve as a reference for other healthcare professionals. Home visits should be initiated by healthcare providers, patients, or family members, with visit frequency and duration adapted to individual needs.

**Conclusions:** This study identified components of a nurse practitioner-led home visit intervention for frail individuals that achieved expert consensus, while highlighting areas where consensus was not reached.

**Clinical Relevance:** These findings will inform the development and future evaluation of such an intervention in real-world settings.

**Highlights:** *What is already known:*

- Frail people living at home have complex clinical, functional, and social needs.
- Home visit interventions may improve access to care for vulnerable populations.
- The content of nurse practitioner-led home visits is not clearly defined.

*What this paper adds:*

- This Delphi study identifies eligibility criteria for nurse practitioner-led home visits, including frailty, multimorbidity, mobility issues, social isolation, and low socioeconomic status.
- It defines key nurse practitioner roles during home visits, such as clinical assessment, treatment adjustment, care coordination, and support for patients and caregivers.
- It specifies modalities of nurse practitioner home visits, including referral pathways, visit duration, and adaptation to patients’ evolving needs.

## INTRODUCTION

Aging at home is the preferred choice for most individuals in the United States (1,2) and Canada (3). The same trend is observed in Europe, where home care remains a priority in many countries (4). However, frailty at any age negatively affects the likelihood of remaining at home (5).

Frailty is defined as a health condition characterized by a decline in functional reserves, making individuals more vulnerable and exposing them to various risks and adverse outcomes (6). Although not exclusive to the geriatric population, frailty is considered a clinical syndrome associated with concerns about the vulnerability and prognosis of older adults and individuals affected by disabling chronic illnesses (7). Multimorbidity, which is particularly prevalent in these populations, further exacerbates frailty (8). Frail patients living at home often experience significant functional limitations that impact their quality of life, a situation that can be improved through home-based interventions (9).

Paradoxically, despite increasing demand (10), the number of home visits is declining worldwide in high income countries (11–14). Yet, home care services have been shown to reduce hospital admissions and nursing home placements (15). Indeed, Coppa et al. (16) reported a 24% reduction in emergency department visits and a 35% decrease in hospital readmissions following the implementation of a home care program. Home care services are also more cost-effective (17,18). For instance, in France, home care costs are three times lower than institutional care (€12,000 vs. €35,000 per person per year) (19).

Nurse practitioners could help address the need for home-based interventions by providing equivalent care to that delivered by physicians, particularly for patients with chronic conditions (20–22). They are also more likely to work in medically underserved and remote areas (23). A nurse practitioner is “a generalist or specialised nurse who has acquired, through additional graduate education (minimum of a master’s degree), the expert knowledge base, complex decision-making skills, and clinical competencies for Advanced Nursing Practice, the characteristics of which are shaped by the context in which they are credentialed to practice.” (24). In the United States, where the role of the nurse practitioner has been well established for several decades, they constitute the bulk of home healthcare providers (25). However, knowledge about the specific nature and extent of their role in home care remains limited (22,26). In Canada, although nurse practitioners have been a part of care for a longer time period, the extent of their role remains a subject of debate (27). In France, nurse practitioners were introduced in 2018 (28), and their role is still being defined within the health system; the situation is similar in Switzerland (29). In Belgium, nurse practitioners have only recently become involved in care services (30). Given the needs of frail adults, a home visit intervention led by a nurse practitioner appears to be a promising solution. Such a program could improve access to care for frail populations and improve healthcare system efficiency. The development of complex healthcare interventions is typically an iterative process involving the identification of needs, the specification of intervention components, and subsequent feasibility testing prior to effectiveness evaluation. The present study represents an early stage of this process by seeking expert consensus on the potential components of a nurse practitioner-led home visit intervention for vulnerable populations.

## Methods

### Aim

The aim of the present study was to consult experts and identify the components of a home visit intervention involving nurse practitioners for vulnerable populations.

### Design

We conducted a consensus study using the Delphi method. The Delphi method is a structured process that leverages expert insights to support decision-making (31). It also enables the anonymous integration of participants with diverse profiles and levels of expertise, preventing any single expert from dominating the consensus process (32).

We employed a modified two-round Delphi method, using an online questionnaire distributed to participants. The two-round Delphi method was chosen over other consensus approaches as it offers greater flexibility and allows a larger number of experts to participate, without requiring them to communicate directly (33). Similarly, the two-round method is designed to determine whether divergent evaluations stem from genuine clinical disagreement (“real” disagreement) or from misunderstandings (“artefactual” disagreement) (33). In line with this approach, the objective of the present study was not to achieve consensus on all items but rather to identify both areas of agreement and areas where expert opinions remained divided. After the second round, items that continued to lack consensus were considered to reflect genuine differences in expert perspectives and were therefore retained as findings of the study rather than subjected to additional rounds.

This study is reported in accord with the CREDES recommendations (Conducting and REporting of DElphi Studies) to ensure methodological rigor and transparency (34). The study was registered with the National Commission for Information Technology and Civil Liberties (CNIL) under the MR-004 reference methodology. In France, in the case of ‘research not involving human subjects’, it is not necessary to consult an ethics committee, provided that a declaration is made to the CNIL. The CNIL declaration number is 2238560 v 0.

### Sample

We recruited a multidisciplinary panel of experts to reflect the diverse specialties involved in treatment decisions (35). Experts were selected through purposive sampling, meaning that only experts from high-income French-speaking countries— Quebec (Canada), Switzerland, Belgium and France—were included. These countries share similar socio-political contexts and healthcare systems, which is necessary for a common perspective on the elements to be selected by consensus (35).

The panel consisted of healthcare professionals from various disciplines (Table 1) who were either practicing home care, working in connection with home care services in rural areas, or engaged in improving access to care for patients with chronic diseases. Experts were recruited through the professional networks of the research team or through official bodies such as professional orders in their respective countries. The first participant was recruited in march 2024. Given that we had previously explored the needs and experiences of frail patients and caregivers living at home through qualitative studies (36,37), patients were not included in the Delphi panel. The present Delphi was not intended to identify patient needs, which had already been explored in prior qualitative work, but rather to define the organisational and clinical components of a future intervention. Consequently, we chose to focus on healthcare professionals with expertise in home care and access to care for vulnerable populations.

**Table 1.** Participant characteristics, n (%)

|  |  |
| --- | --- |
| Female sex | 10 (66.7) |
| *Practice location* |  |
| Quebec | 4 (26.7) |
| Switzerland | 3 (20.0) |
| Belgium | 3 (20.0) |
| France | 5 (33.3) |
| *Occupation* |  |
| General practitioner | 7 (46.7) |
| Nurse Practitioner | 7 (46.7) |
| Other (occupational therapist) | 1 (6.6) |
| *Type of practice experience* |  |
| Rural area | 7 (46.7) |
| Remote area | 3 (20.0) |
| Home care | 5 (33.3) |

### Data collection

The questionnaire was developed using the French version of the Template for Intervention Description and Replication (TIDieR) checklist (Appendix A) (38) and the needs of patients and caregivers identified in a prior focus group (36). The primary aim of the TIDieR checklist is to encourage authors to describe interventions in sufficient detail to ensure their replicability (38). It emphasizes key aspects of an intervention, including its purpose and content (why, what), the individuals involved (who), the mode of delivery (how), the timing (when), and the duration (how long). Additionally, the checklist accounts for adaptations based on user needs as the intervention is implemented.

Although the TIDieR checklist was originally developed as a reporting tool, its domains were used in the present study to ensure that key intervention characteristics were systematically considered during the identification of potential intervention components.

The initial questionnaire comprised 12 questions, to which experts responded using a 9-point rating scale (where 1 indicated complete disagreement and 9 indicated complete agreement) (35,39). The questionnaire also included open-ended text fields labelled “other suggestions” to further the discussion in the second round. Open-ended questions generate ideas and provide more flexibility in participant responses making it possible to identify key issues for the next round (Table 3) (40). Additionally, sociodemographic data were collected at the end of the questionnaire, including domain of expertise, gender, and age group, last degree completed, country and location of practice. The final version was reviewed and approved by the steering committee of the doctoral thesis of which this study formed part.

For each round, the questionnaire was distributed online via Sphinx Online (Le Sphinx, Chavanod, France) to all participating experts. The second round consisted of a revised questionnaire including only six questions with five items requiring further consensus, and ten new items based on suggestions provided in the “other suggestions” section (35). Participants also received feedback on the first-round results along with the second-round questionnaire. Each submission-response cycle corresponded to one Delphi round (31). The first round was conducted in May–June 2024, followed by the second round in August–September 2024.

### Ethical considerations

This study was registered with the National Commission for Information Technology and Civil Liberties (CNIL) under the MR-004 reference methodology, which applies to research not involving human participants but conducted in the health field. The CNIL declaration number is 2238560 v 0. All participants received comprehensive information to ensure their free and informed consent regarding their participation, as well as the objectives of the study and its potential publication. Consent was obtained in writing at multiple stages, including during the initial contact and at each round of the study.

### Data analysis

For each questionnaire item, we calculated the median score to determine whether the item should be retained. Consensus was defined as a median score ≥7, with no disagreement among experts.

Disagreement was defined as one-third or more of participants rating an item within both extreme ends of the 9-point scale (1–3 or 7–9). Items were considered rejected by consensus if the median score was ≤3. Items that did not reach consensus in the first round were included in the second round of the Delphi process.

The study was not registered.

## Results

### Participant characteristics

A total of 19 experts meeting the eligibility criteria were contacted. The majority of experts took part in both rounds of the Delphi survey (first round n = 15; 79% and second round n = 14; 74%).

Among the participants, 10 (66.7%) were women. There was an equal proportion of nurse practitioners (n = 7; 47%) and family physicians (n = 7; 47%). The participants’ characteristics are detailed in Table 1.

### Selected items

Of the 61 items presented in the first round, 51 (83.6%) reached consensus, while five (8.2%) were rejected. Another five items (8.2%) did not reach consensus and were carried forward to the second round. Additionally, 10 new items were added based on comments provided by seven professionals (46.7%) from different countries and professions during the first round. This resulted in a total of 15 items being reassessed in the second round (Table 3). Among these, eight (53.3%) were accepted, while seven (46.7%) did not reach consensus.

Items that reached final consensus were used to define the *potential target populations* for a nurse practitioner-led home care intervention. Experts agreed that this intervention should focus primarily on older adults, as well as individuals with physical and/or cognitive disabilities or those receiving end-of-life care. Regarding the *patients’ eligibility criteria*, key factors highlighted by experts included the Rockwood Frailty Scale (scoring>5) (41). The presence of multimorbidity, mobility issues, social isolation (defined in the questionnaire as a lack of a support network), and low socioeconomic status, defined with the European Deprivation Index (EDI) (42).

The *role of the nurse practitioner* providing home care and the *key parameters that should be assessed for appropriate patient management* were also selected (Table 2). Experts emphasized the importance of clinical assessment, adjusting treatments, and ensuring the continuity of care. They also highlighted the need for nurse practitioners to be able to communicate whenever needed with family members and with the following healthcare and social services providers and entities: home care providers, social workers, rehabilitation professionals, administrative services (e.g. : Departmental center for disabled persons, National Health Insurance) hospital services (e.g., emergency services, post-acute and rehabilitation care, general care), health and social services organizations. Additionally, experts agreed that nurse practitioners should interact with the family physician in case of emergency, changes in the patient’s condition, or whenever deemed necessary by either the nurse practitioner or the family physician. However, they also acknowledged that family physicians should continue conducting home visits when required.

**Table 2.** Nurse practitioner in Home Visits: Key Responsibilities and Evaluation Metrics.

| **Roles of the nurse practitioner at home** | **Parameters to Evaluate for Adequate Care** |
| --- | --- |
| Clinical assessment | Cognitive function |
| Adjusting treatments | Functional abilities |
| Continuity of care | Frailty index |
| Coordination of the care team | Professionals involved |
| Resource for the patient | Treatment adherence |
| Referral to specialized resources | Available support from relatives |
| Adaptation of care objectives and life goals | Care satisfaction |
| Prevention and health promotion | Pain management |
| Therapeutic education and self-care promotion | Nutritional status |
|  | Screening for abuse |
|  | Achievement of care objectives |
|  | Caregiver burnout level |

**Table 3.**
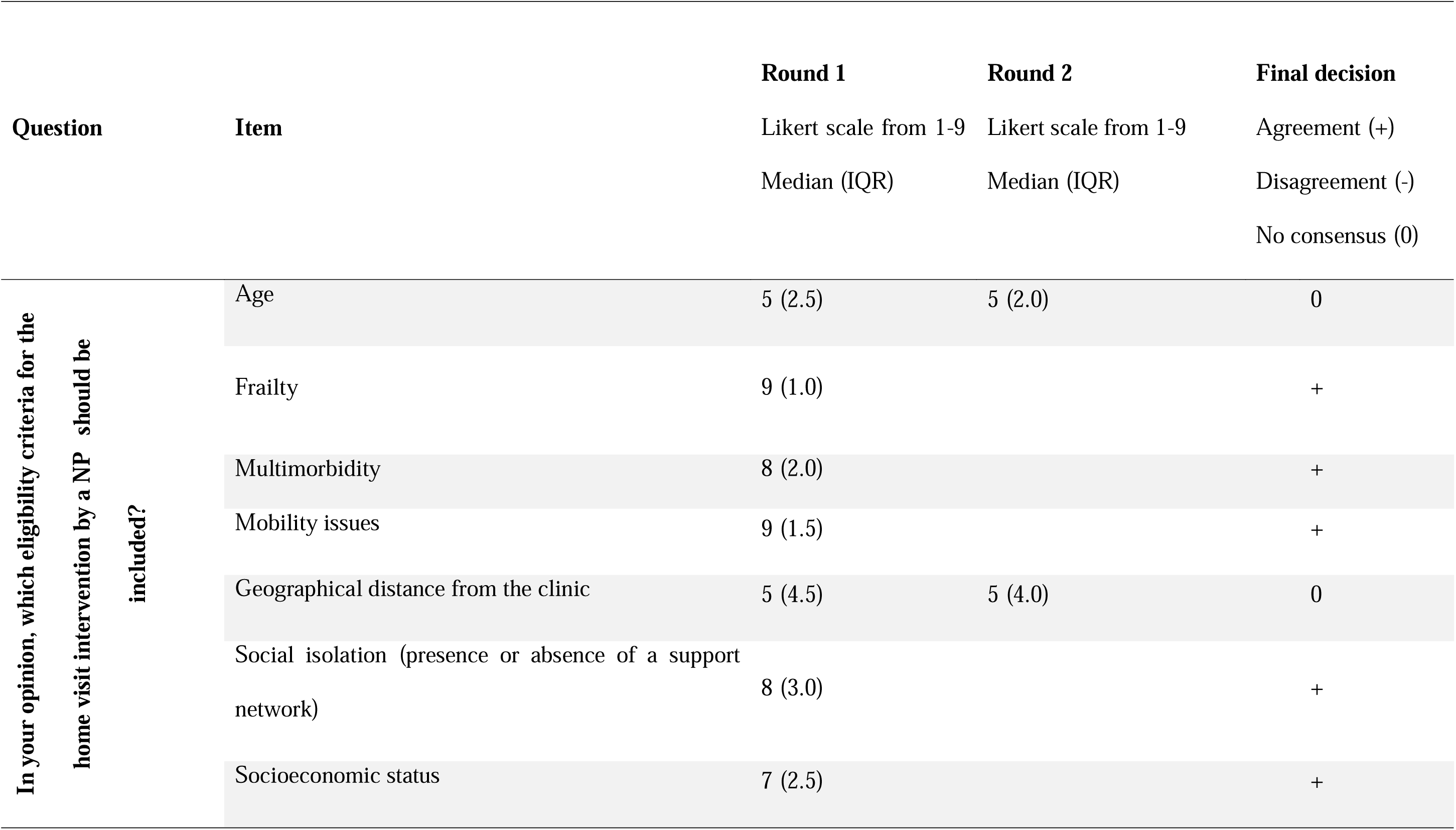
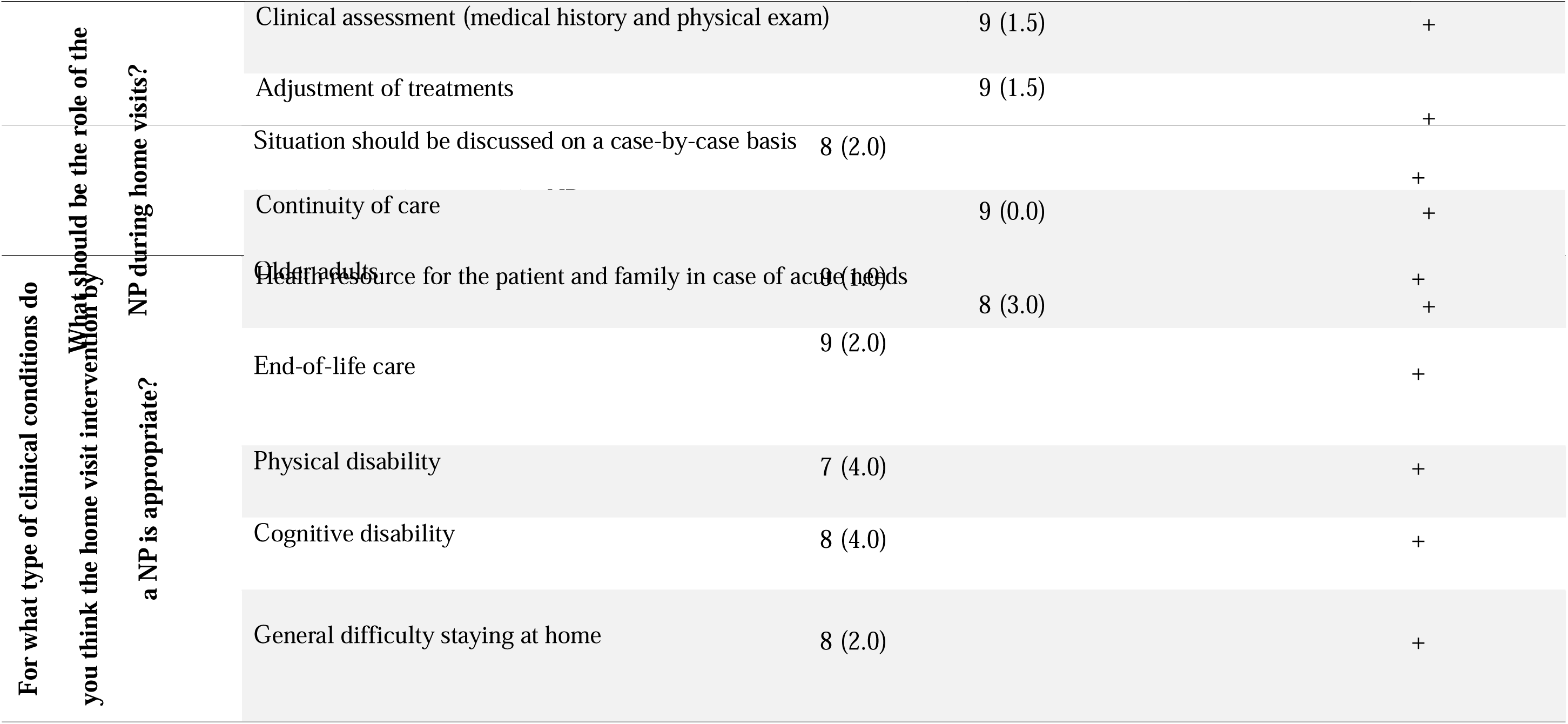
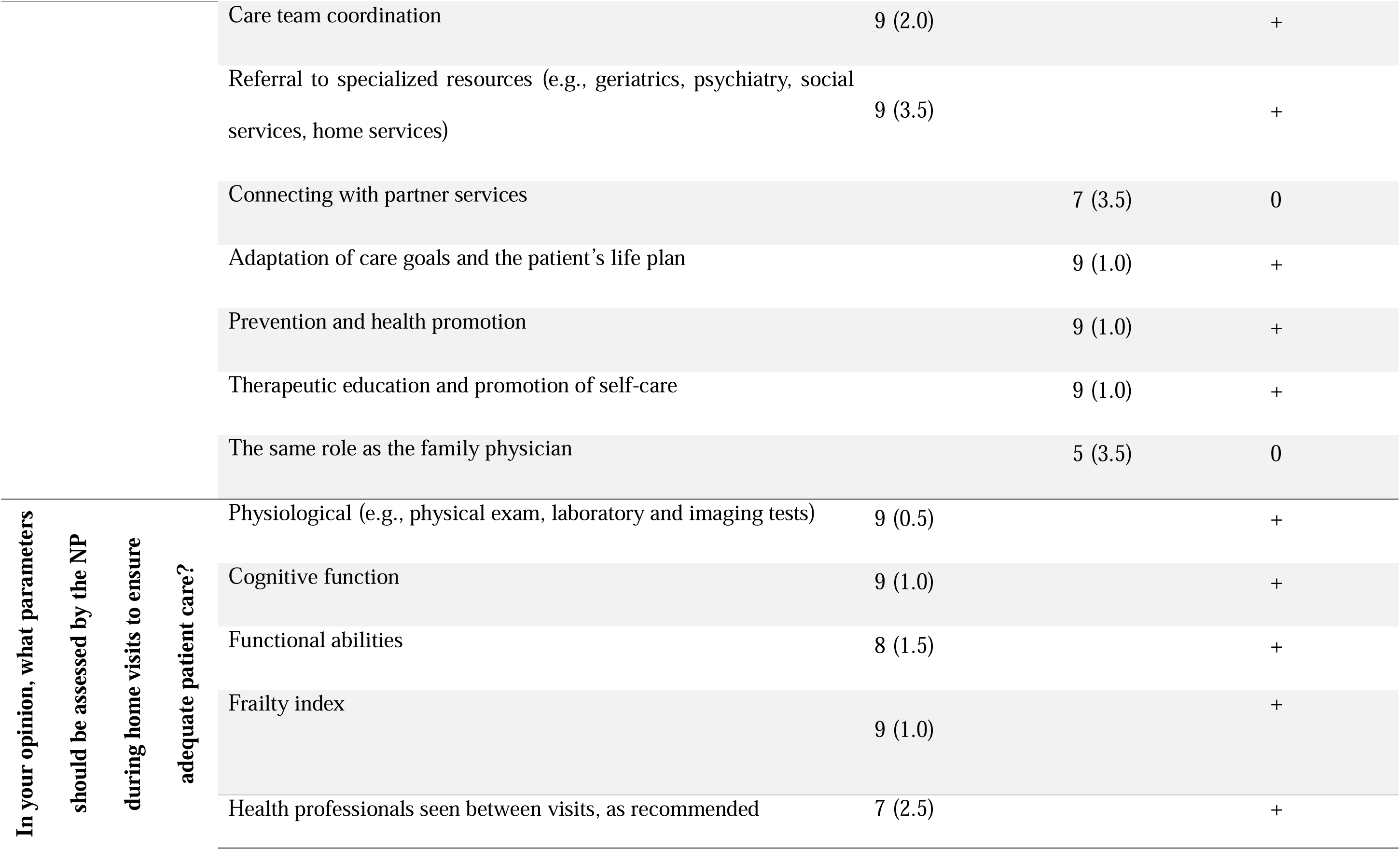
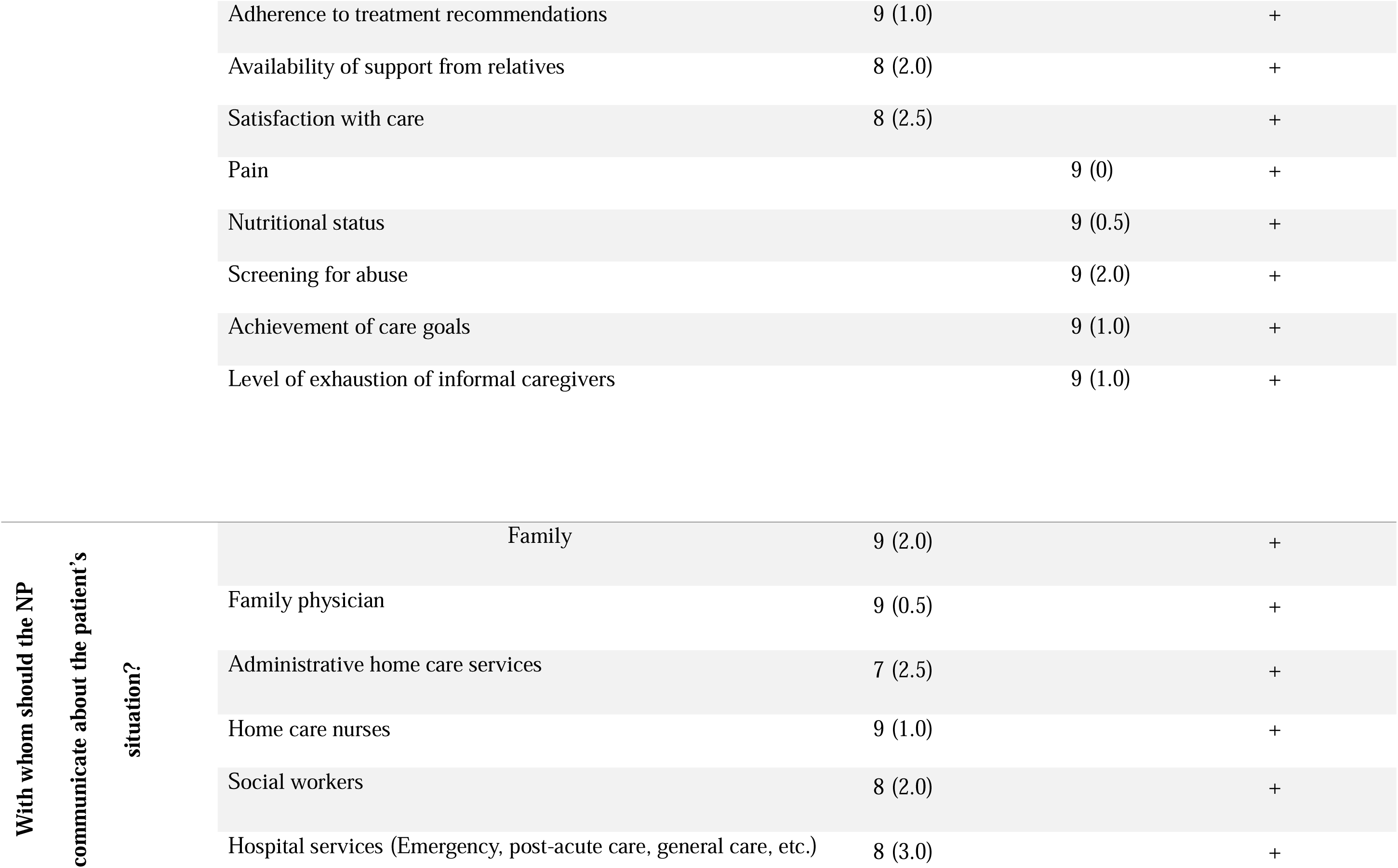
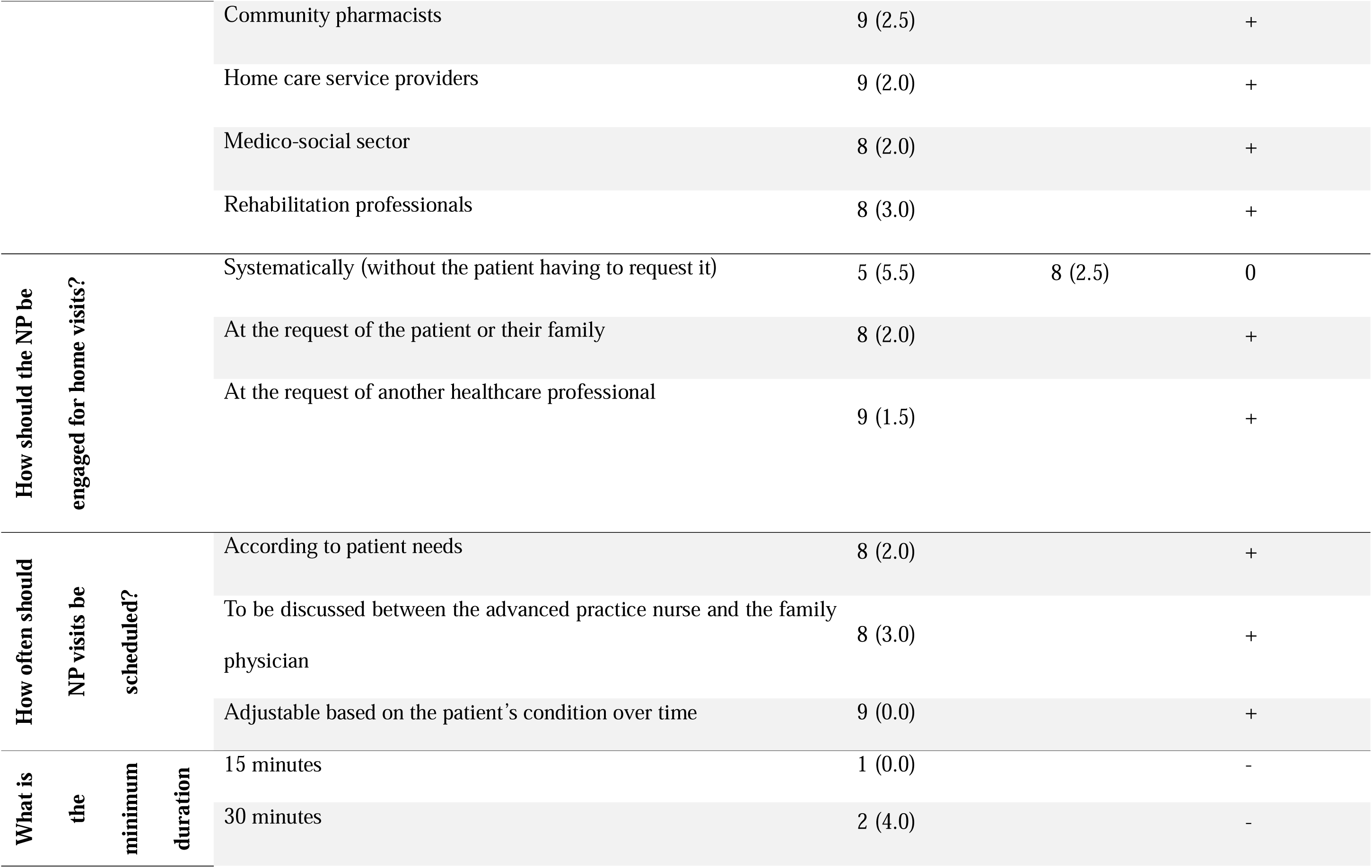
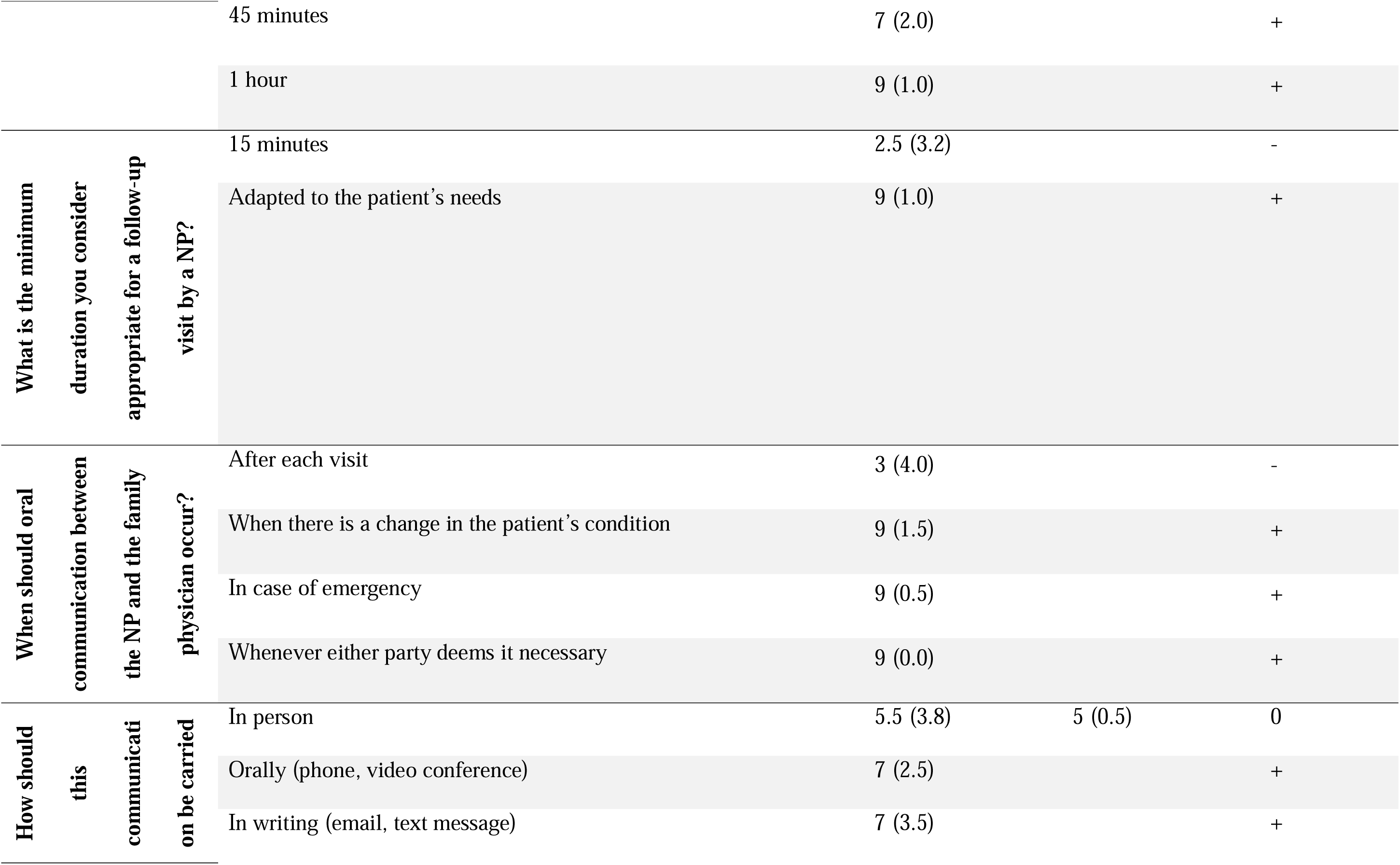
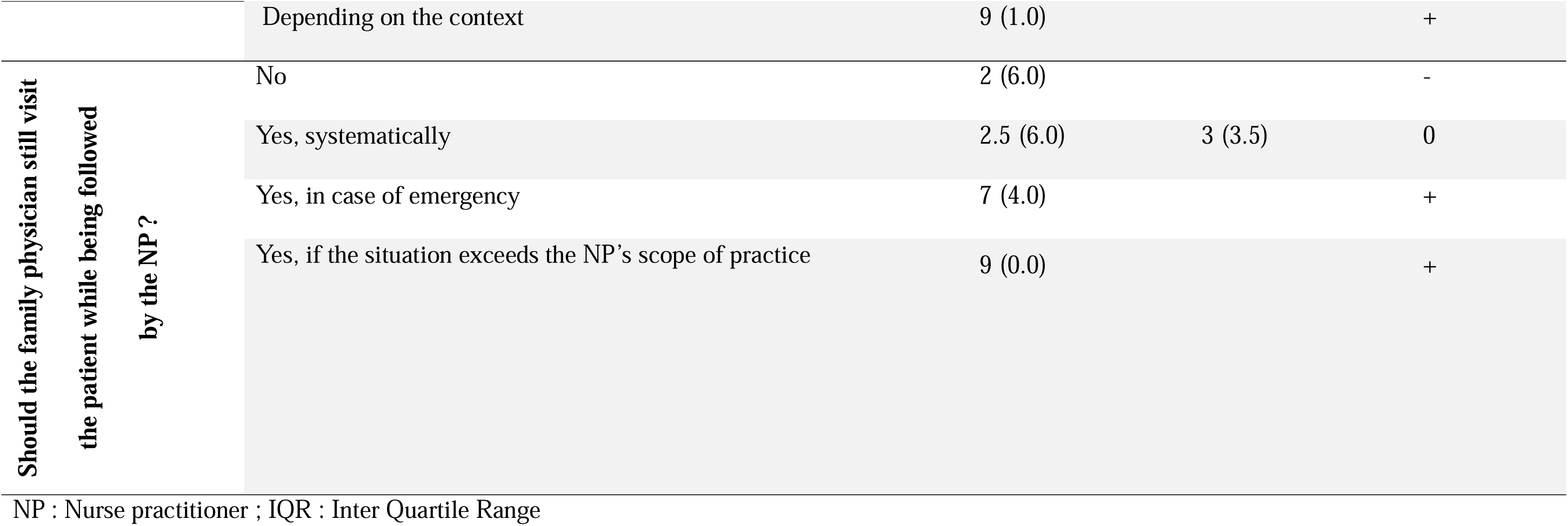
Delphi round’s results.

Regarding the *administration of the intervention*, experts agreed that the nurse practitioners should become involved at the request of another healthcare professional, the patient, or their family, provided that the patient meets the eligibility criteria. Additionally, the intervention should take place at the home of eligible patients, regardless of the geographical distance from the family medicine clinic. Concerning the *timing and duration of the intervention*, experts agreed that patient’s needs should guide the frequency of home visits, following a discussion between the nurse practitioners and the family physician regarding the required clinical follow-up, and should remain adaptable as the patient’s condition evolves. A consensus was also established for a minimum duration of 45 minutes to one hour for the initial visit, while follow-up visits should last at least 15 minutes and be adjusted according to the patient’s needs.

Seven items did not reach consensus. Experts were divided on two eligibility criteria: age and geographical distance from the family medicine clinic. Experts also disagreed on whether establishing connections with various healthcare professionals, referred to as “partners,” should be considered part of the nurse practitioner’s role. Similarly, experts did not agree on whether the nurse practitioner’s role should be equivalent to that of the family physician. Communications could be transmitted either orally or in writing, the experts did not have a preference, and the modality had to be adapted to the context. Additionally, experts were divided on whether a home visit by the physician should still be mandatory if the nurse practitioner was already providing care at home.

## Discussion

This study aimed to identify the key components of a nurse practitioner-led home care intervention in remote areas. To achieve this, international experts, mostly nurse practitioners and family physicians, involved in home care were consulted. Experts defined the target populations for the intervention as: older adults, individuals with physical or cognitive disabilities, receiving end-of-life care, or experiencing challenges to stay at home. Regarding the role of the nurse practitioner, experts agreed that it should encompass both clinical care and coordination of the healthcare network, acting as the patient’s primary point of reference. They also emphasized that the frequency and duration of visits should be tailored to the specific needs of each patient.

These eligibility criteria align with findings from a systematic review on the implementation of home visiting programs led by nurses (43), although the nurses in the review were not nurse practitioners. The review highlighted that functional autonomy, including physical and mental impairment, nutritional status, mobility issues, and social isolation, were commonly used criteria in most home visiting programs. While frailty assessment was another eligibility criterion, the studies relied on scales other than the one developed by Rockwood (41). However, the systematic review (43) identified a minimum age criterion (60 years), which was rejected by the experts in our study.

Although age as an eligibility criterion did not reach consensus, experts acknowledged that the intervention would be appropriate for both older patients and other frail individuals. While age is correlated with frailty, it is not its sole predictive factor (41). The accumulation of deficits, unintentional weight loss, chronic exhaustion, multimorbidity, mental health, lifestyle factors, and social vulnerability were also identified as significant determinants of frailty (41). This aligns with the criteria our experts identified for nurse practitioners to assess in home care patients.

Social isolation was also considered an eligibility criterion in the present study. In rural areas, we might expect to find more individuals who experience greater isolation. However, research by Henning-Smith et al. (44) and Bonnell et al. (45) indicates that social isolation is actually more prevalent in urban settings. Consequently, there was no consensus among experts regarding the inclusion of geographical distance as a criterion. Similarly, Cudjoe et al. (46), reported that geographical distance between a family medicine clinic and a patient’s home is not correlated with isolation, although it may influence it. Indeed, their study found no significant association between the home’s geographic location and the risk of social isolation (OR = 1.21, 95% CI 0.93-1.57, non-significant) (46). However, where people live can still shape their social interactions in indirect ways. Rural areas, for example, tend to have fewer social resources, different household compositions due to younger generations moving to cities, and lower income and education levels, all of which can contribute to social isolation (46).

While experts debated the role played by geographical distance in isolation, another consideration was whether telehealth could compensate for the lack of home visits and reduce costs (47). While telehealth follow-up can enhance accessibility, in-person visits remain crucial for comprehensive assessments and patient engagement (48). Older adults face significant barriers to digital technology use, with only 47% of French people aged 75 and more owning a computer, and access further limited by socioeconomic disparities (49). Replacing home visits with telehealth alone carries the risk of exacerbating inequalities, making a hybrid approach the most effective solution (48).

The scoping review by De Leede-Brunsveld et al. (50) described the pivotal role played by nurse practitioners in patient management, providing care coordination while also serving as a clinical expert, which aligns with the findings of the present study. The International Council of Nurses (ICN) also recognizes the nurse practitioner’s role as a care coordinator and primary point of contact for patients and families, as stated in the Guidelines on Advanced Nursing Practice (2020). Research findings and recognized organizations thus acknowledge the nurse practitioner as a key figure in patient management, working as a team with the family physician, which the experts in this study also agreed upon.

However, experts disagreed on whether establishing connections with other healthcare providers (e.g., home care nurses, social workers, hospital services, community pharmacists) should be part of the nurse practitioner’s role. Studies on nurse practitioner’s responsibilities in Europe report that she/he can assume this function as part of their care coordination role (50). The distinction between care coordination and establishing connections with other healthcare providers is subtle. Therefore, despite the lack of consensus on this specific aspect, this function should not be excluded from the nurse practitioner’s role in home interventions.Regarding visit frequency, a literature review indicates that it is often determined based on a needs assessment or according to a pre-established but flexible schedule (51). Research highlights the benefits of individualized care, a person-centered approach, adherence to recommendations, and multiple visits (52). Nurse practitioners adopt a more person-centered approach, focusing on comprehensive and coordinated care, being more responsive to the needs of vulnerable patients, and fostering greater patient and family involvement (53).

The idea that nurse practitioners could substitute for family physicians generated disagreement among experts. This may stem from the controversial status of the nurse practitioner role in Canada (27) and France (54), its uncertain definition in Switzerland (29), and the fact that its legal framework has yet to be established in French-speaking Belgium, despite the presence of training programs (55,56). The lack of expert consensus could also reflect a desire to differentiate between nurse practitioners and physicians, to allow each profession to define its unique contributions more clearly. Positioning nurse practitioners as equivalent to physicians implies parity in skills and effectiveness, whereas defining a distinct role for the nurse practitioner emphasizes the unique contribution their nursing expertise can bring, and the added value of combining two different perspectives (57,58). The longstanding opposition between the medical and nursing cultures (59) is often framed through the lens of “care” versus “cure.” The nursing profession is traditionally associated with humanization and “caring,” while medicine is linked to technical expertise and “curing.” However, rather than being in opposition, these two approaches complement each other perfectly in a collaborative healthcare model (60). Recognizing the distinct yet complementary nature of nurse practitioners and physicians may be key to enhancing interprofessional collaboration and improving patient care.

### Study Strengths and Limitations

This study is characterized by a high participation rate, reflecting a keen interest in this topic. Expert feedback was considered to refine the questionnaire for the second round, ensuring that it closely aligned with the opinions put forth on nurse practitioner-led home visits. The panel included experts from both medical and paramedical backgrounds, knowledgeable in home care in rural, semi-urban, and remote areas, across four different countries. This diversity increases the likelihood that key factors relevant to a nurse practitioner-led home intervention for patients with multimorbidity were accurately identified. Additionally, the study provided an international perspective, enhancing the feasibility of developing an intervention that can be adapted to different healthcare systems.

While a third Delphi round might have resulted in additional consensus, the persistence of disagreement after feedback and reassessment suggests that these items reflected genuine differences in expert opinion rather than ambiguity in questionnaire wording.

Although the participation rate was satisfactory, the number of experts consulted was limited. However, this study represents the first step toward developing an intervention aimed at optimizing home care for frail patients by nurse practitioners. The next phase will involve co-constructing the intervention with multiple stakeholders—including healthcare professionals, clinical decision-makers, and patients/families—to ensure that necessary adjustments are made to align with the actual needs and preferences of the target population. In addition, despite selecting countries with a shared language and comparable socioeconomic levels, cultural and healthcare system differences may have influenced expert responses and contributed to certain disagreements, particularly regarding the definition of the nurse practitioner’s role. Therefore, a future intervention will need to account for contextual elements specific to each country or healthcare region to ensure its acceptability, feasibility and effectiveness.

Finally, this Delphi study should be viewed as an initial step in the development of a complex intervention. Consistent with recommendations for complex intervention development, the present work aimed to identify and refine potential intervention components before proceeding to stakeholder co-design, feasibility assessment, and future effectiveness evaluation.

## Conclusion

This study has highlighted the key determinants of a nurse practitioner-led home visit program for frail patients. Eligibility criteria for such an intervention include frailty assessment, the presence of multimorbidity, mobility issues, social isolation, and low socioeconomic status. The nurse practitioner’s role should cover clinical assessment, adjusting treatments, care coordination, therapeutic education, self-care promotion, and support for patients and their families. Additionally, the nurse practitioner could serve as a primary point of contact for other healthcare providers. Nurse practitioner-led home visit could be initiated at the request of a healthcare professional, the patient, or a family member, and should be tailored in terms of frequency and duration to meet individual needs. Findings from this study will provide a foundation to develop an intervention aimed at improving home-based are for frail patients, which will be followed by a feasibility assessment to make the required adjustments prior to conducting a full pragmatic trial.

## Data Availability

All data produced in the present study are available upon reasonable request to the authors

## Declarations

## Ethics approval and consent to participate

This study was conducted in accordance with the ethical principles outlined in the Declaration of Helsinki. Approval from an institutional ethics committee could not be obtained for this project, since ethics committees in France only evaluate projects involving the collection of clinical or population data, or those conducted as part of a clinical trial. However, in compliance with France legal requirements for research, the study was registered with the National Commission for Information Technology and Civil Liberties (CNIL) under the MR-004 reference methodology (no. 2238560). Similar to institutional ethics committees, the CNIL ensures that data privacy laws are applied to the collection, storage and use of personal data, while also ensuring that human rights are respected. All participants received comprehensive information to ensure their free and informed consent regarding their participation, as well as the objectives of the study and its potential publication. Written consent was obtained at multiple stages, including during the initial contact and at each round of the study. Completing the Delphi questionnaire constituted implicit consent to participate in the study. Approval from an ethics committee was therefore not required in the various countries where participants were recruited.

## Consent for publication

Not applicable

## Authors’ contributions

MB designed the study. AS collected the data. AS and AB analyzed the data. AS and CP prepared the manuscript. All authors approved the final version for submission. All listed authors meet the authorship criteria and confirm their agreement with the content of the manuscript. This research was conducted in France.

## Acknowledgments

The authors would like to sincerely thank all the experts who participated in the Delphi survey. Their generous contribution of time and expertise was essential to the success of this study. the authors acknowledge the Asalée Association for supporting the doctoral research from which this article is derived.

## Declaration of generative AI and AI-assisted technologies in the manuscript preparation process

Generative AI and AI-assisted tools were not used to generate, modify, or enhance any figures, images, graphical abstracts, or artwork in this manuscript.

## Appendix

## Appendix A. TiDier Checklist

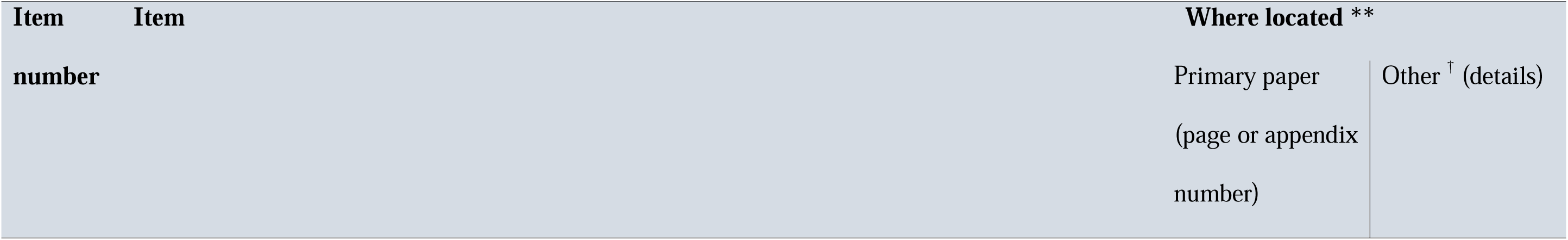
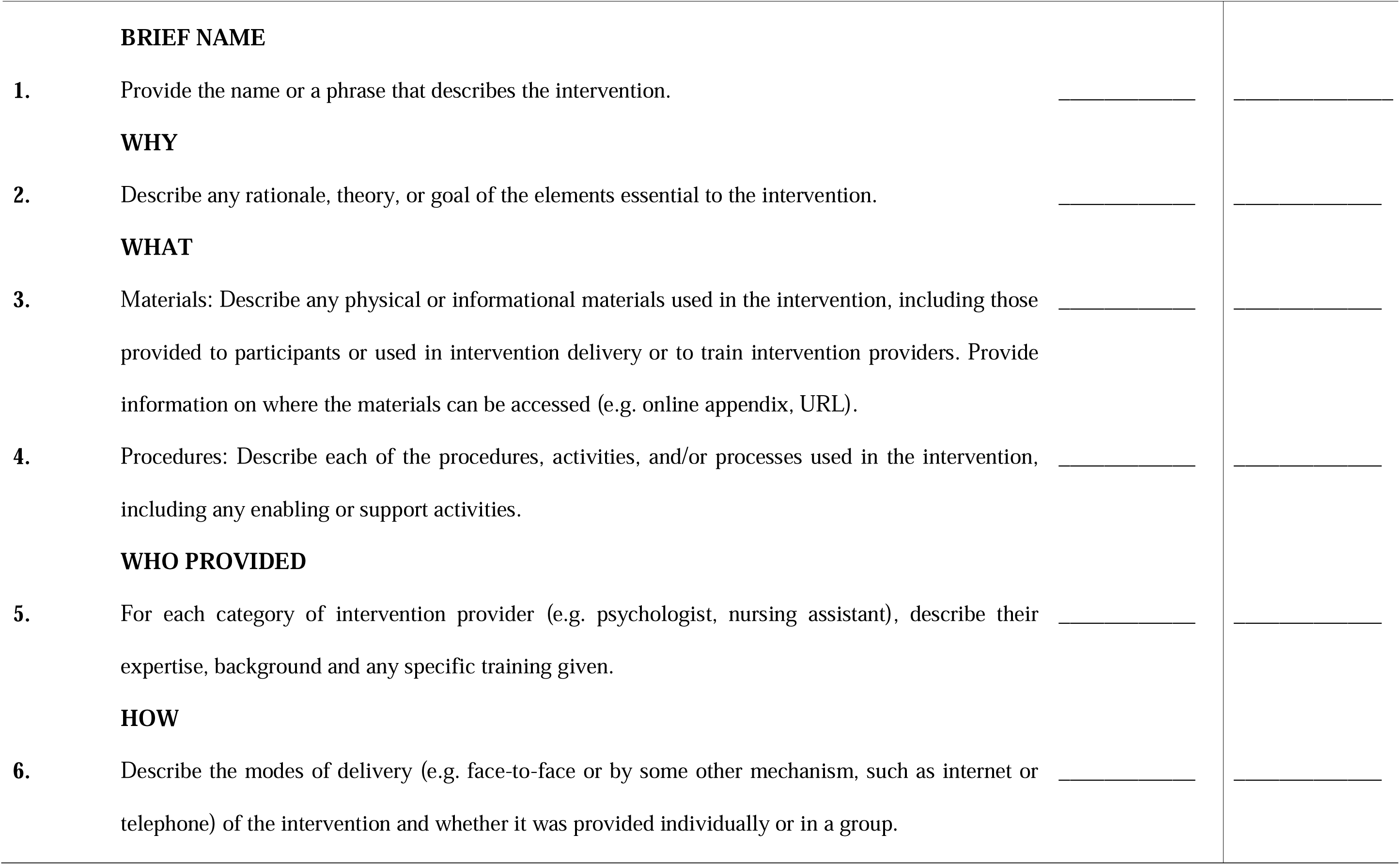
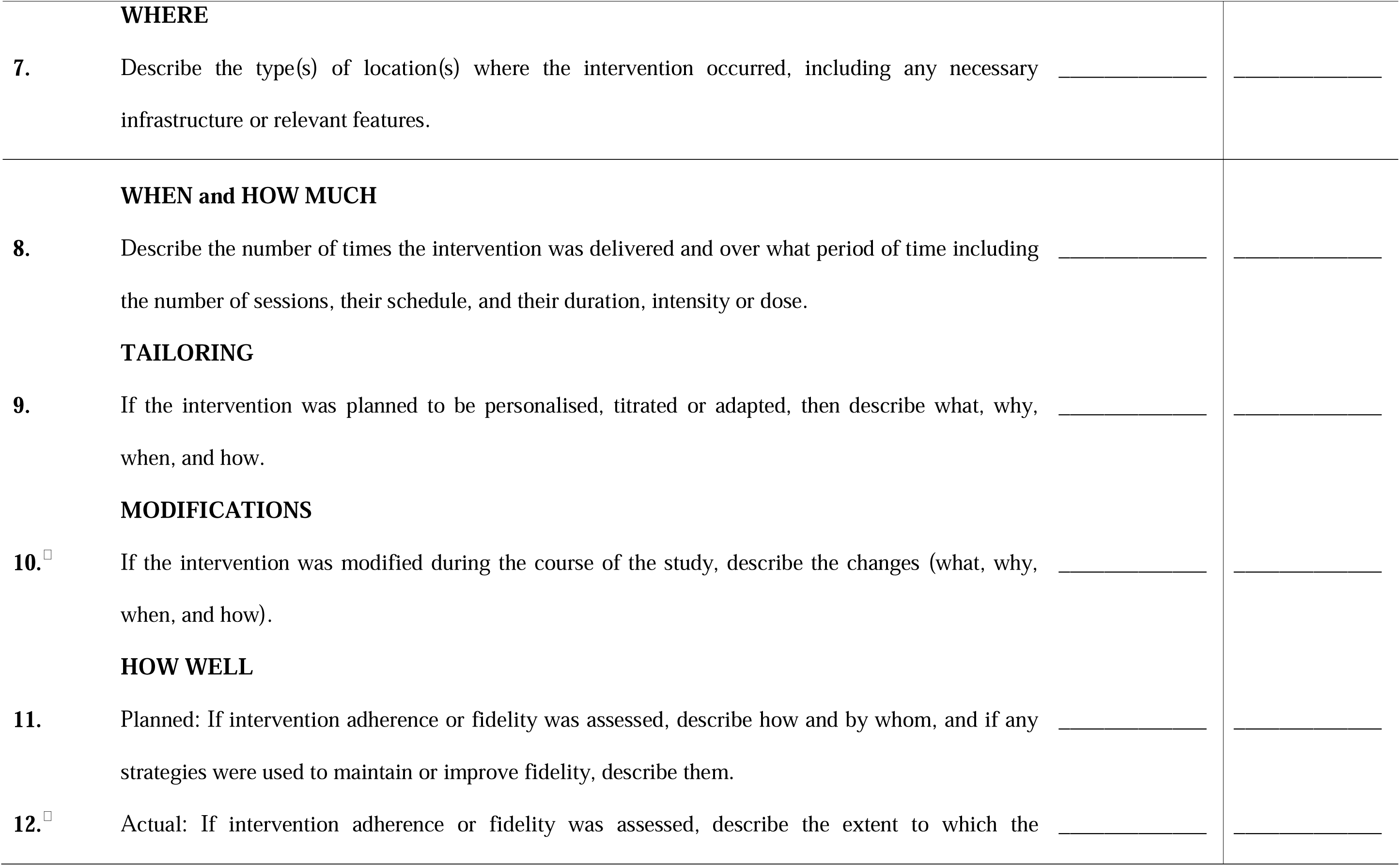
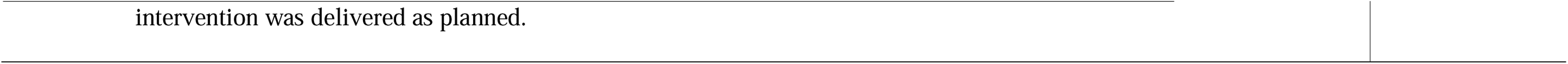

